## Supplemental Materials for "Longitudinal analyses after COVID-19 recovery or prolonged infection reveal unique immunological signatures after repeated vaccinations"

Daisuke Hisamatsu et al.

### Supplemental Materials

A

| | | WT | WT | WT | Mutants |
|---|---|---|---|---|---|
| **Severity** | **Healthy** | **Mild** | **Moderate I** | **Moderate II** | **Moderate II** |
| Number of paticipants | 5 | 10 | 10 | 10 | 8 |
| Age (mean ± SD, years) | 41.4 ± 17.3 | 36.3 ± 17.3 | 63.7 ± 14.6 | 70.5 ± 11.3 | 52.6 ± 7.3 |
| Sex (% male) | 40 | 70 | 60 | 60 | 75 |
| Time from symptom onset to bood collection on admission (mean ± SD, days) | NA | 2.9 ± 1.7 | 5.8 ± 1.2 | 8.3 ± 4.1 | 8.0 ± 2.8 |
| Time from admission to discharge (mean ± SD, days) | NA | 7.8 ± 5.6 | 11.1 ± 2.4 | 14.5 ± 6.3 | 18.3 ± 5.0 |
| **Clinical factors on admission** | | | | | |
| SpO2 (mean ± SD, %) | NA | 97.8 ± 1.4 | 96.7 ± 1.4 | 90.9 ± 4.7 | 94.3 ± 1.7 |
| Fibrinogen (mean ± SD, mg/dL) | NA | 317.7 ± 55.8 | 486.4 ± 102.5 | 662.0 ± 160.8 | 533.1 ± 115.0 |
| Ferritin (mean ± SD, ng/mL) | NA | 169.0 ± 108.8 | 629.0 ± 475.2 | 680 ± 449.2 | 932.6 ± 300.0 |
| D-dimer (mean ± SD, µg/mL) | NA | 1.6 ± 0.4 | 1.6 ± 0.3 | 2.5 ± 0.8 | 1.5 ± 0.2 |
| IL-6 (mean ± SD, pg/mL) | NA | 3.6 ± 2.6 | 22.1 ± 18.3 | 42.4 ± 32.6 | 29.3 ± 11.2 |
| Krebs von den Lungen-6(KL-6) (mean ± SD, U/mL) | NA | 238.0 ± 144.5 | 336.9 ± 469.1 | 260.0 ± 104.5 | 256.1 ± 109.7 |
| Serum Amyloid A (SAA) (mean ± SD, µg/mL) | NA | 17.6 ± 18.7 | 386.0 ± 428.5 | 2029.5 ± 1804.3 | 346.2 ± 262.7 |
| pulmonary Surfactant Protein-D (SP-D) (mean ± SD, ng/mL) | NA | 24.6 ± 8.5 | 36.1 ± 45.3 | 57.5 ± 70.4 | 34.5 ± 41.8 |
| White Bood Cell (WBC) (mean ± SD, $x10^3$/µl) | NA | 5.2 ± 1.8 | 4.9 ± 1.9 | 8.0 ± 3.2 | 4.9 ± 2.3 |
| Hemoglobin (Hb) (mean ± SD, g/dL) | NA | 15.1 ± 1.8 | 13.7 ± 1.5 | 13.3 ± 0.5 | 15.0 ± 0.8 |
| Neutrophil (NE) (mean ± SD, %) | NA | 59.9 ± 11.5 | 68.7 ± 9.8 | 81.5 ± 11.3 | 71.5 ± 9.7 |
| Lymphocyte (Lym) (mean ± SD, %) | NA | 28.2 ± 9.0 | 22.7 ± 8.8 | 11.8 ± 8.8 | 20.1 ± 7.7 |
| Monocyte (Mo) (mean ± SD, %) | NA | 8.7 ± 3.1 | 7.7 ± 2.4 | 5.3 ± 3.2 | 7.8 ± 2.5 |
| Eosinophil (Eos) (mean ± SD, %) | NA | 2.8 ± 2.7 | 0.7 ± 0.8 | 0.2 ± 0.3 | 0.1 ± 0.2 |
| C-Reavtive Protein (CRP) (mean ± SD, mg/dL) | NA | 0.3 ± 0.1 | 4.4 ± 5.3 | 11.6 ± 7.2 | 5.0 ± 3.5 |

B

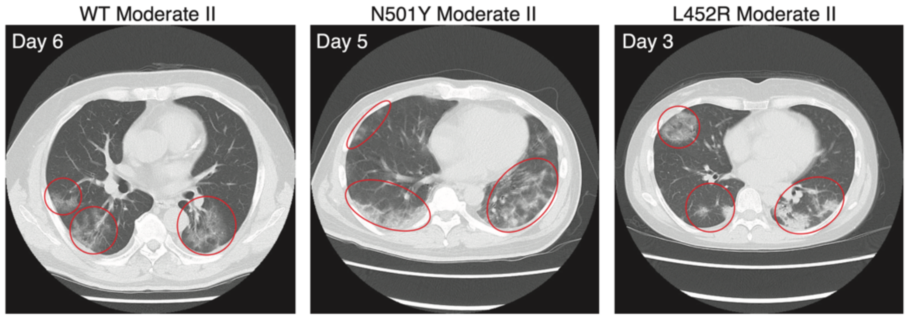


**Supplemental Figure 1**

**Supplemental Figure 1. Study cohort.** (A) The table shows clinical information of all participants in this study. (B) Representative images of COVID-19 pneumonia in moderate II patients.

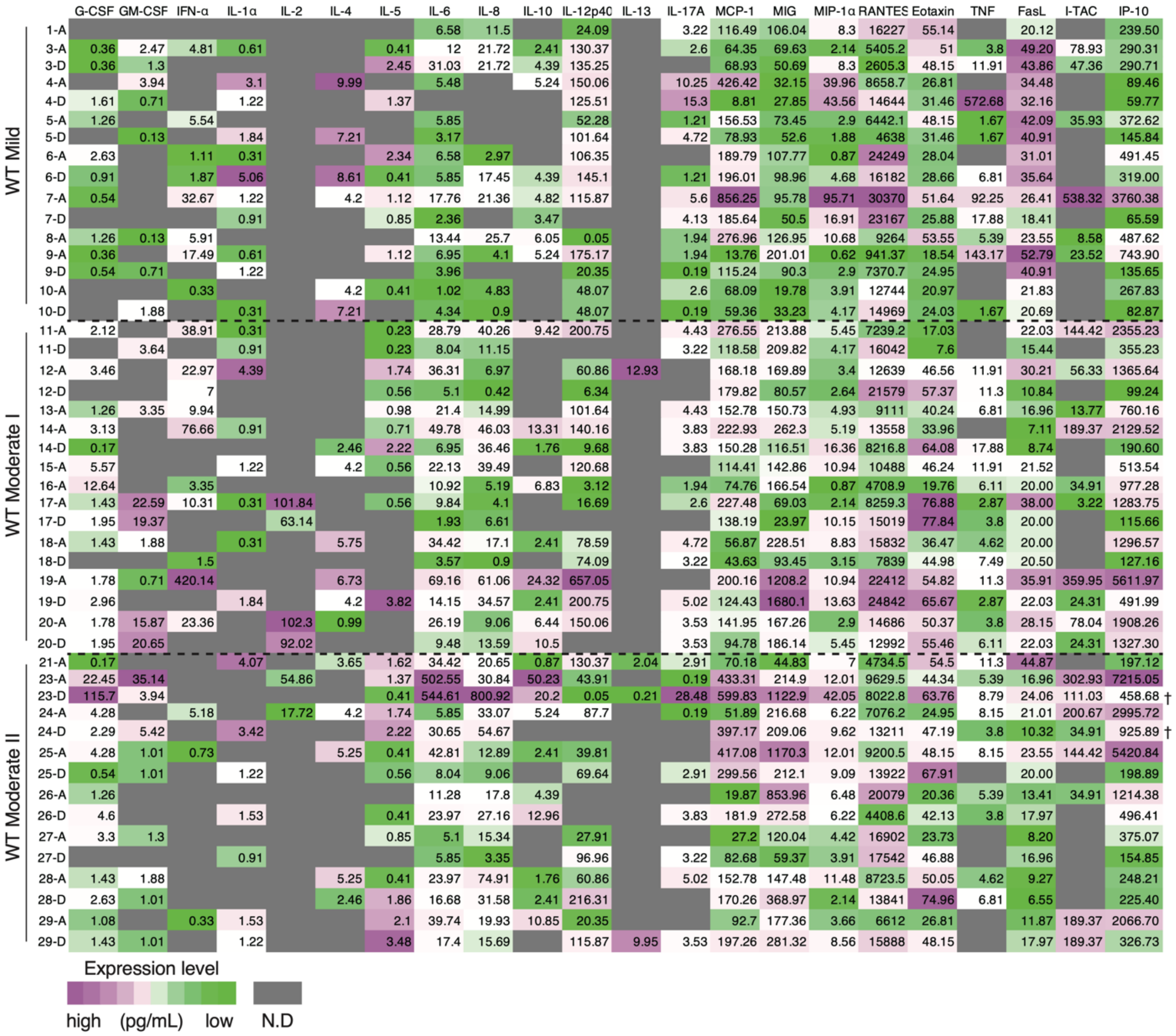


**Supplemental Figure 2**

**Supplemental Figure 2. Comprehensive cytokine expression analysis in COVID-19 patients infected with WT.** Patient identification numbers are shown. A indicates admission and D indicates discharge.

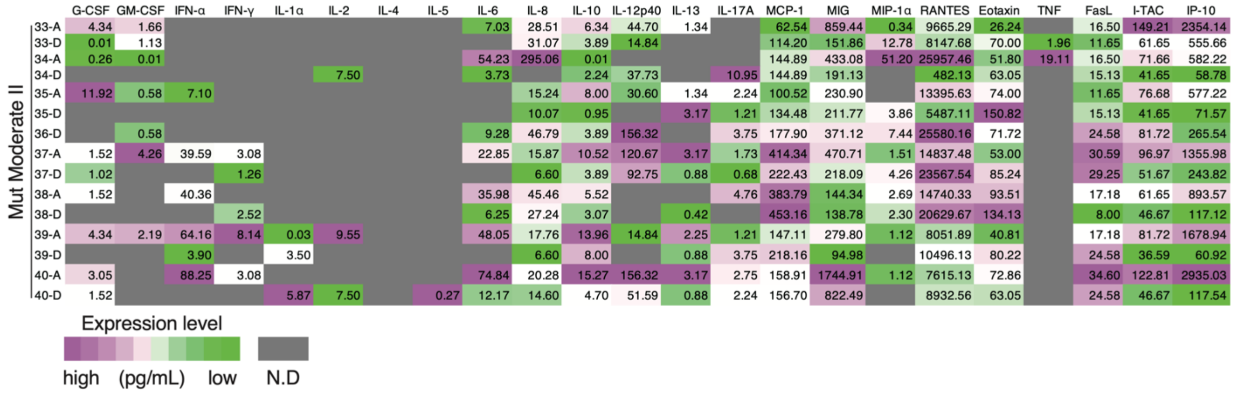


| | G-CSF | GM-CSF | IFN-α | IFN-γ | IL-1α | IL-2 | IL-4 | IL-5 | IL-6 | IL-8 | IL-10 | IL-12p40 | IL-13 | IL-17A | MCP-1 | MIG | MIP-1α | RANTES | Eotaxin | TNF | FasL | I-TAC | IP-10 |
|---|---|---|---|---|---|---|---|---|---|---|---|---|---|---|---|---|---|---|---|---|---|---|---|
| 33-A | 4.34 | 1.66 | | | | | | | 7.03 | 28.51 | 6.34 | 44.70 | 1.34 | | 62.54 | 859.44 | 0.34 | 9665.29 | 26.24 | | 16.50 | 149.21 | 2354.14 |
| 33-D | 0.01 | 1.13 | | | | | | | | 31.07 | 3.89 | 14.84 | | | 114.20 | 151.86 | 12.78 | 8147.68 | 70.00 | 1.96 | 11.65 | 61.65 | 555.66 |
| 34-A | 0.26 | 0.01 | | | | | | | 54.23 | 295.06 | 0.01 | | | | 144.89 | 433.08 | 51.20 | 25957.46 | 51.80 | 19.11 | 16.50 | 71.66 | 582.22 |
| 34-D | | | | | | 7.50 | | | 3.73 | | 2.24 | 37.73 | | 10.95 | 144.89 | 191.13 | | 482.13 | 63.05 | | 15.13 | 41.65 | 58.78 |
| 35-A | 11.92 | 0.58 | 7.10 | | | | | | | 15.24 | 8.00 | 30.60 | 1.34 | 2.24 | 100.52 | 230.90 | | 13395.63 | 74.00 | | 11.65 | 76.68 | 577.22 |
| 35-D | | | | | | | | | | 10.07 | 0.95 | | 3.17 | 1.21 | 134.48 | 211.77 | 3.86 | 5487.11 | 150.82 | | 15.13 | 41.65 | 71.57 |
| 36-D | | 0.58 | | | | | | | 9.28 | 46.79 | 3.89 | 156.32 | | 3.75 | 177.90 | 371.12 | 7.44 | 25580.16 | 71.72 | | 24.58 | 81.72 | 265.54 |
| 37-A | 1.52 | 4.26 | 39.59 | 3.08 | | | | | 22.85 | 15.87 | 10.52 | 120.67 | 3.17 | 1.73 | 414.34 | 470.71 | 1.51 | 14837.48 | 53.00 | | 30.59 | 96.97 | 1355.98 |
| 37-D | 1.02 | | | 1.26 | | | | | | 6.60 | 3.89 | 92.75 | 0.88 | 0.68 | 222.43 | 218.09 | 4.26 | 23567.54 | 85.24 | | 29.25 | 51.67 | 243.82 |
| 38-A | 1.52 | | 40.36 | | | | | | 35.98 | 45.46 | 5.52 | | | 4.76 | 383.79 | 144.34 | 2.69 | 14740.33 | 93.51 | | 17.18 | 61.65 | 893.57 |
| 38-D | | | | 2.52 | | | | | 6.25 | 27.24 | 3.07 | | 0.42 | | 453.16 | 138.78 | 2.30 | 20629.67 | 134.13 | | 8.00 | 46.67 | 117.12 |
| 39-A | 4.34 | 2.19 | 64.16 | 8.14 | 0.03 | 9.55 | | | 48.05 | 17.76 | 13.96 | 14.84 | 2.25 | 1.21 | 147.11 | 279.80 | 1.12 | 8051.89 | 40.81 | | 17.18 | 81.72 | 1678.94 |
| 39-D | | | 3.90 | | 3.50 | | | | | 6.60 | 8.00 | | 0.88 | 3.75 | 218.16 | 94.98 | | 10496.13 | 80.22 | | 24.58 | 36.59 | 60.92 |
| 40-A | 3.05 | | 88.25 | 3.08 | | | | | 74.84 | 20.28 | 15.27 | 156.32 | 3.17 | 2.75 | 158.91 | 1744.91 | 1.12 | 7615.13 | 72.86 | | 34.60 | 122.81 | 2935.03 |
| 40-D | 1.52 | | | | 5.87 | 7.50 | | 0.27 | 12.17 | 14.60 | 4.70 | 51.59 | 0.88 | 2.24 | 156.70 | 822.49 | | 8932.56 | 63.05 | | 24.58 | 46.67 | 117.54 |

**Supplemental Figure 3**

**Supplemental Figure 3. Comprehensive cytokine expression analysis in COVID-19 patients infected with VOCs.** Patient identification numbers are shown. A indicates admission, and D indicates discharge.

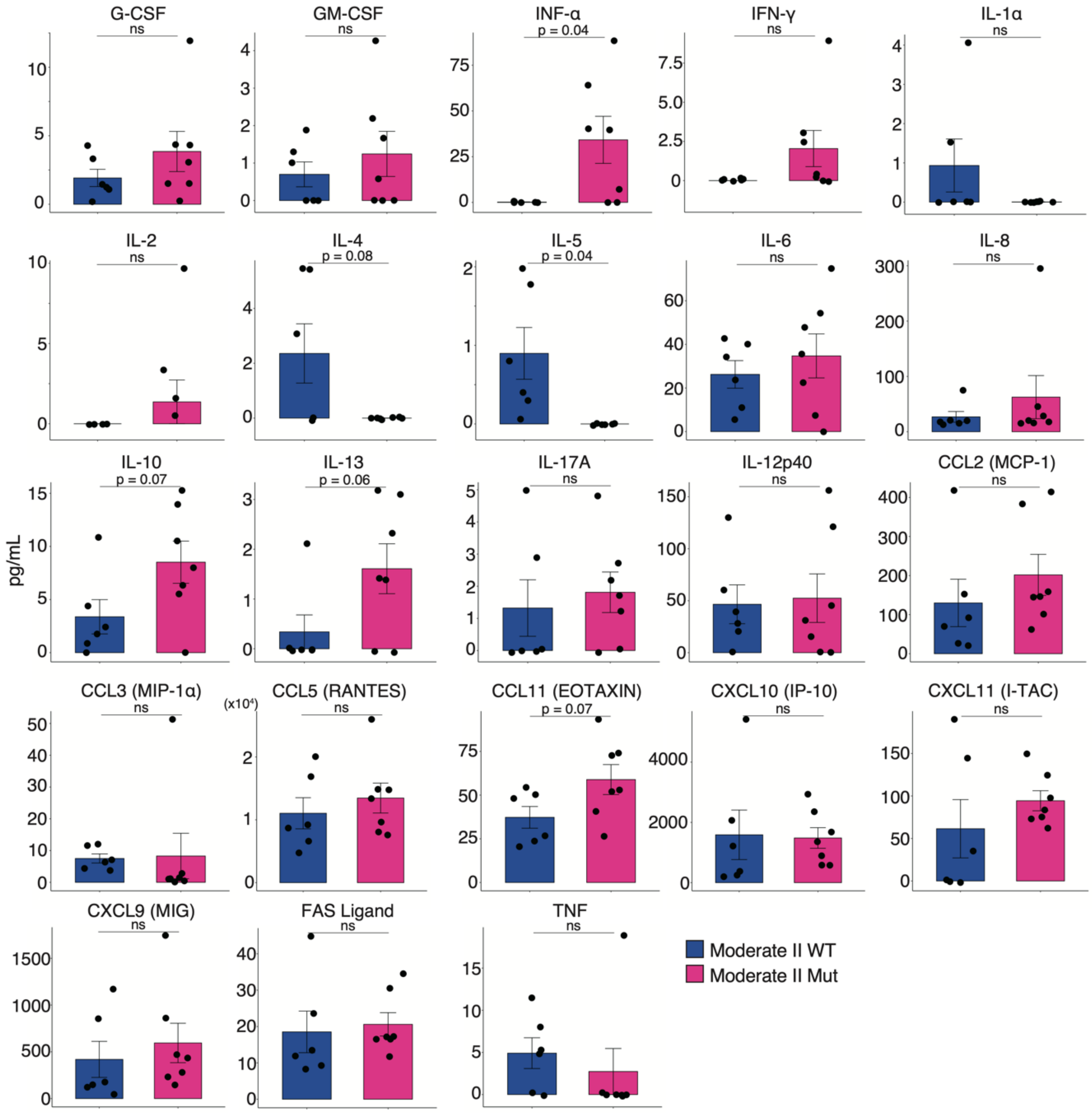


**Supplemental Figure 4**

**Supplemental Figure 4. Comparison of cytokine expressions in moderate II patients infected with the WT strain or VOCs on admission.** Graphs show the concentration of each cytokine. Statistical significance was determined using Welch's t-test. ns, not significant.

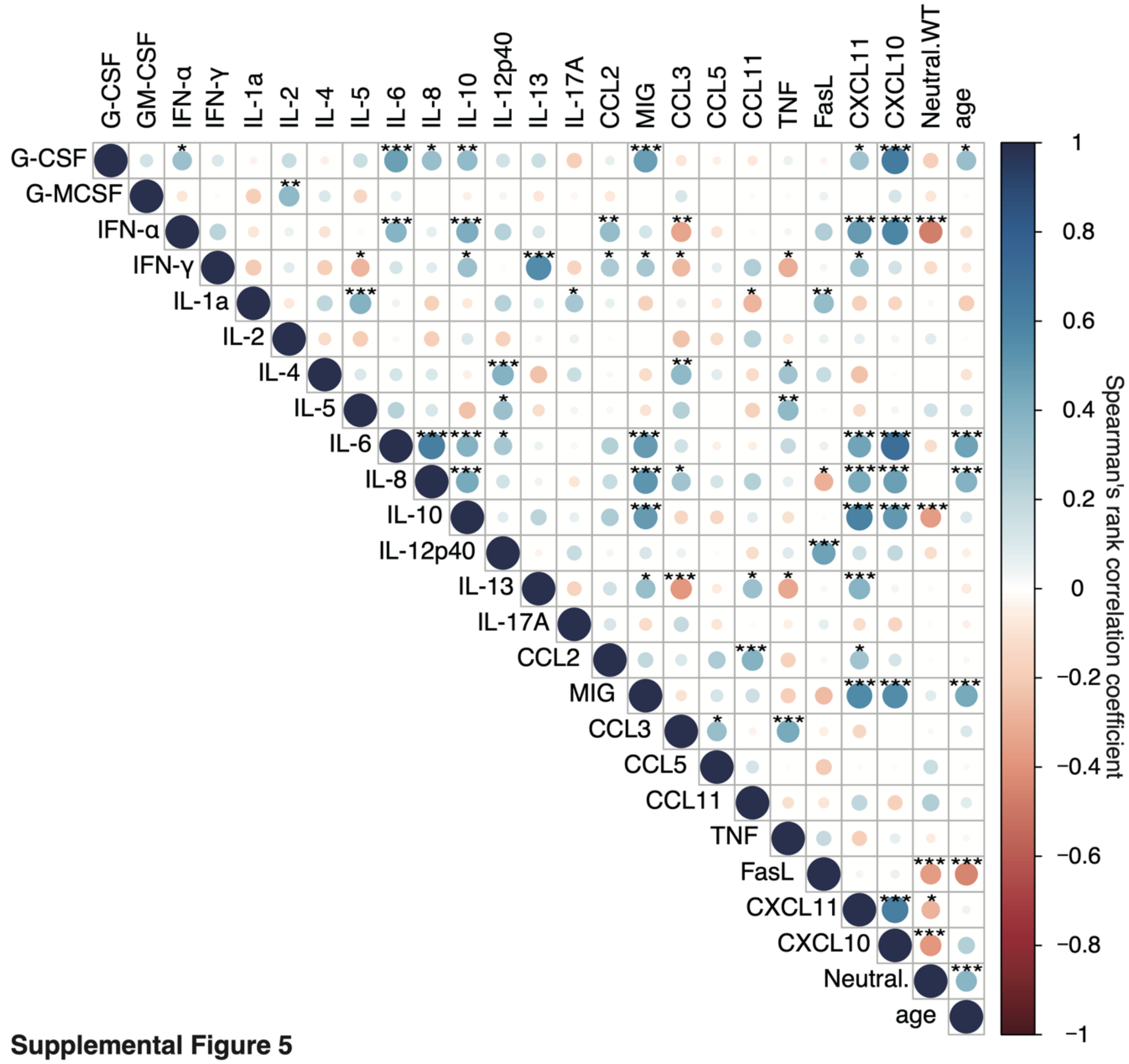


**Supplemental Figure 5**

**Supplemental Figure 5. Correlation analysis among cytokine expression, neutralizing activity, and age in COVID-19 patients during hospitalization period.** Correlation matrices were created using the Spearman's correlation coefficient. *p < 0.05; **p < 0.01; ***p < 0.005.

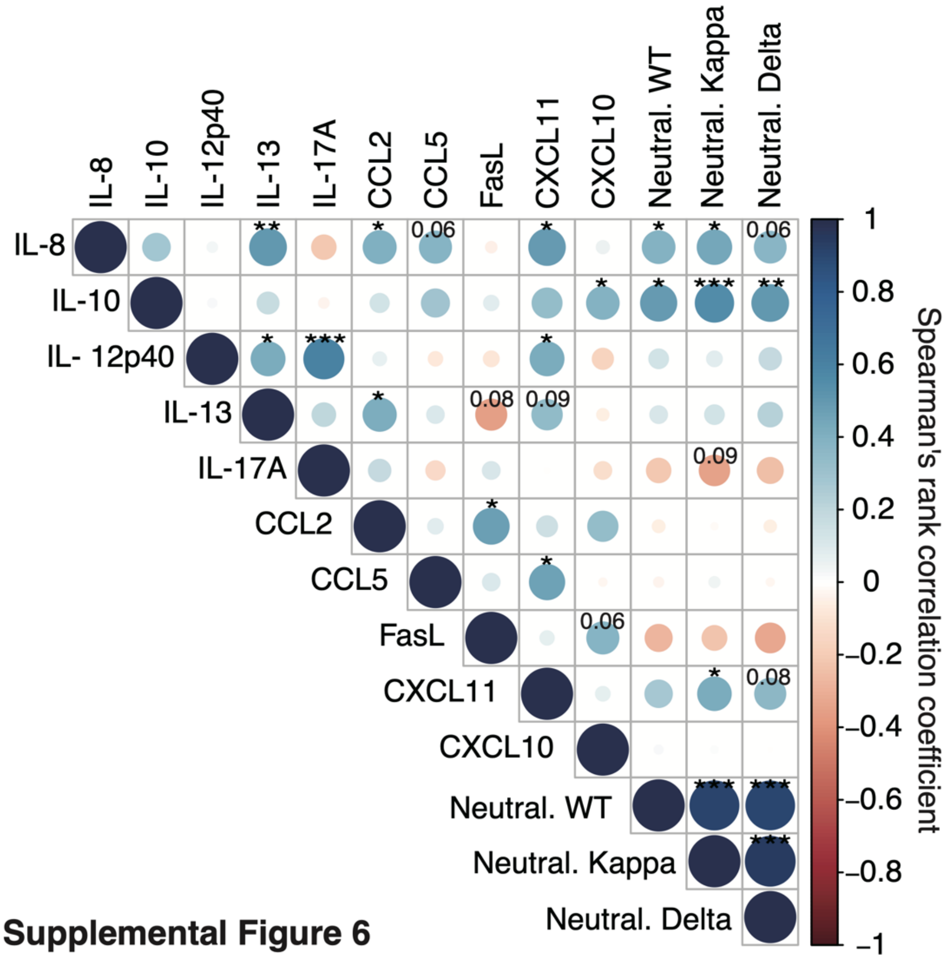


**Supplemental Figure 6**

**Supplemental Figure 6. Correlation analysis of cytokine expression and neutralizing activity in all participants, including naïve and recovered individuals.** Correlation matrices were created using the Spearman's correlation coefficient. *$p < 0.05$; **$p < 0.01$; ***$p < 0.005$.

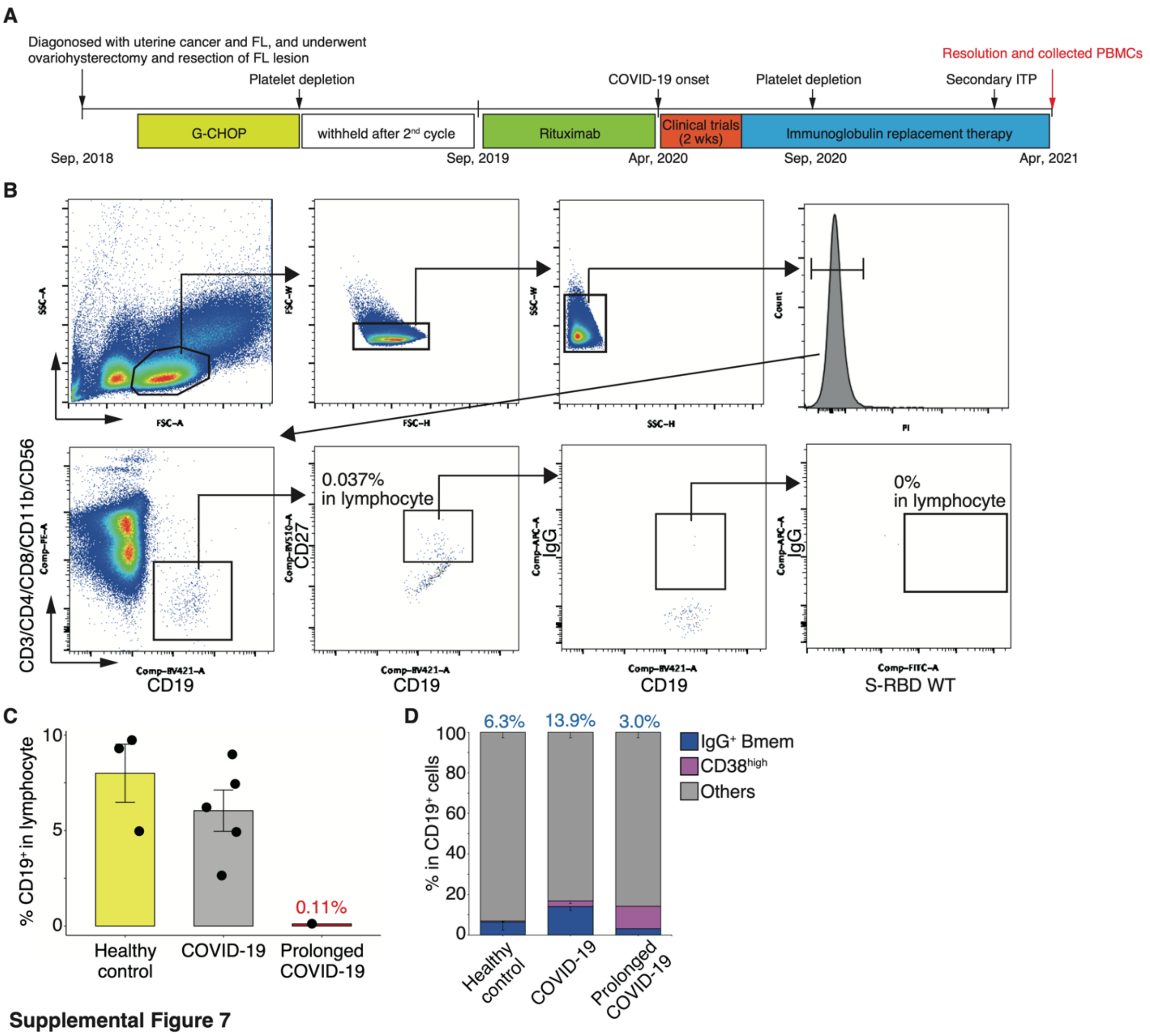


**Supplemental Figure 7**

**Supplemental Figure 7. Prolonged SARS-CoV-2 infection in a patient with follicular lymphoma undergoing B-cell depletion therapy.** (A) Graph showing the clinical course and treatment strategy. FL, follicular lymphoma; ITP, immune thrombocytopenia; G-CHOP, obinutuzumab plus cyclophosphamide, doxorubicin, vincristine, and prednisone therapy. (B) Representative FACS plots of the cell population are shown in panels. The number indicates the positive rate. (C) The graph shows the proportion of $CD19^+$ cells in each group. (D) Graph showing the frequencies of $IgG^+$ Bmems and $CD38^{high}$ cell populations. Each number indicates the mean value.

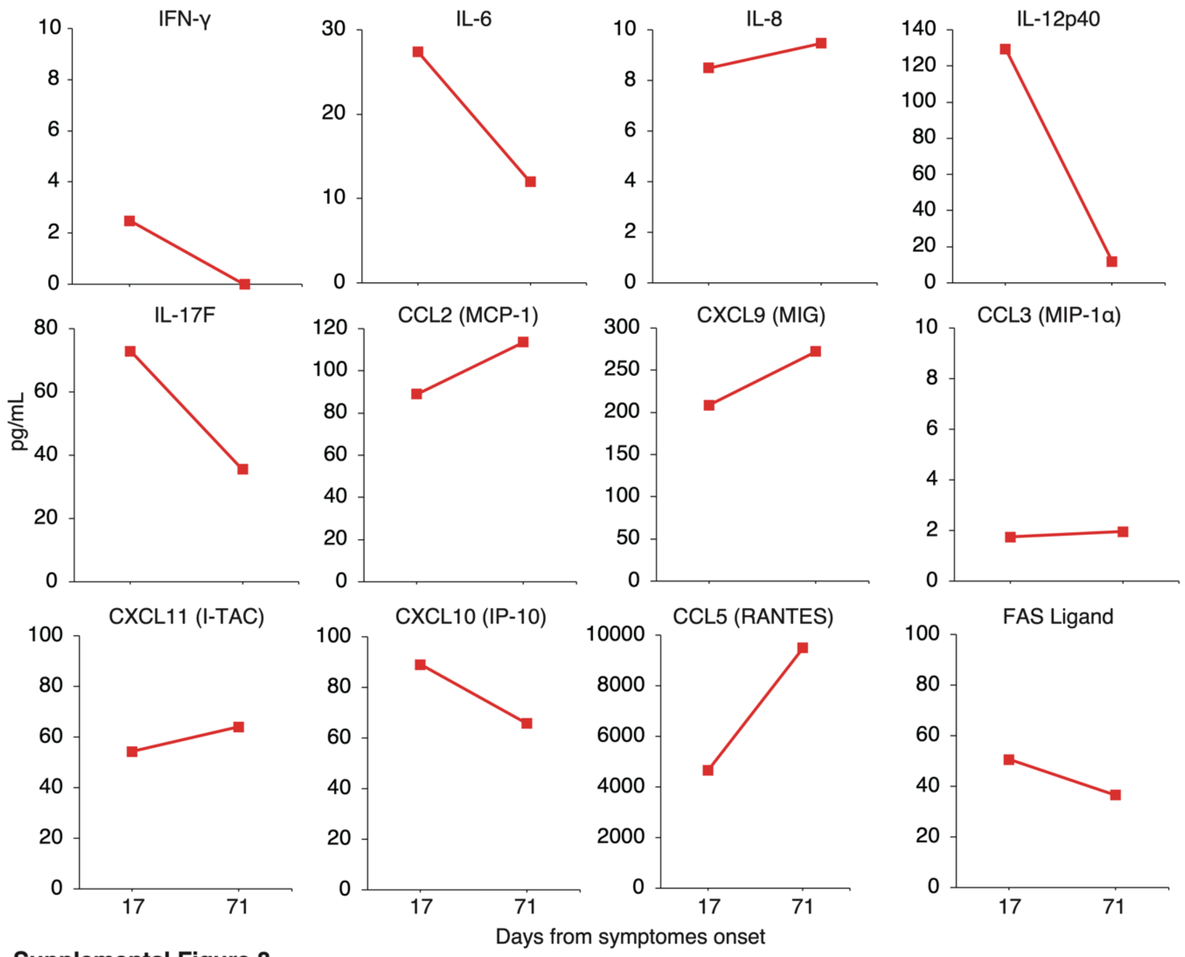


**Supplemental Figure 8**

**Supplemental Figure 8. Cytokine expression change in a patient with prolonged COVID-19.**

Graphs show representative cytokine concentrations.

**Forward Primer**

| Target | Sequence |
|---|---|
| IGH_variable region (v) 1 | 5-ACAGGTGCCCACTCCCAGGTGCAG-3 |
| IGH_v3 | 5-AAGGTGTCCAGTGTGARGTGCAG-3 |
| IGH_v4/6 | 5-CCCAGATGGGTCCTGTCCCAGGTGCAG-3 |
| IGH_v5 | 5-CAAGGAGTCTGTTCCGAGGTGCAG-3 |
| IGL(κ)_v1/2 | 5-ATGAGGSTCCCYGCTCAGCTGCTGG-3 |
| IGL(κ)_v3 | 5-CTCTTCCTCCTGCTACTCTGGCTCCCAG-3 |
| IGL(κ)_v4 | 5-ATTTCTCTGTTGCTCTGGATCTCTG-3 |
| IGL(λ)_v1 | 5-GGTCCTGGGCCCAGTCTGTGCTG-3 |
| IGL(λ)_v2 | 5-GGTCCTGGGCCCAGTCTGCCCTG-3 |
| IGL(λ)_v3 | 5-GCTCTGTGACCTCCTATGAGCTG-3 |
| IGL(λ)_v4/5 | 5-GGTCTCTCTCSCAGCYTGTGCTG-3 |
| IGL(λ)_v6 | 5-GTTCTTGGGCCAATTTTATGCTG-3 |
| IGL(λ)_v7 | 5-GGTCCAATTCYCAGGCTGTGGTG-3 |
| IGL(λ)_v8 | 5-GAGTGGATTCTCAGACTGTGGTG-3 |

**Reverse Primer**

| Target | Sequence |
|---|---|
| IGH_constant region (c) 1 | 5-CGCCTGAGTTCCACGACACC-3 |
| IGH_c for nested PCR | 5-TCGGGGAAGTAGTCCTTGAC-3 |
| IGL(κ)_c1 | 5-GAGGCAGTTCCAGATTTCAA-3 |
| IGL(κ)_c for nested PCR | 5-GGGAAGATGAAGACAGATGGT-3 |
| IGL(λ)_c1 | 5-GCTTGAAGCTCCTCAGAGG-3 |
| IGL(λ)_c for nested PCR | 5-GGGCGGGAACAGAGTGACC-3 |

**Supplemental Table**

**Supplemental Table. Primer sets of single-cell RT-PCR and immunoglobulin gene sequencing**
